# Low dose oral ketamine treatment on post-traumatic stress disorder (PTSD) (OKTOP): An open-label pilot study

**DOI:** 10.1101/2024.11.26.24318024

**Authors:** Bonnie L. Quigley, Adem T. Can, Megan Dutton, Cyrana C. Gallay, Grace Forsyth, Monique Jones, Fiona Randall, Trish Wilson, Jim Lagopoulos, Daniel F. Hermens

## Abstract

Ketamine is being actively investigated as a rapid-acting treatment for many conditions with a stress-related psychopathology, including post-traumatic stress disorder (PTSD). The majority of studies regarding ketamine treatment for PTSD to date (including open-label and randomised control trials) have focused on intravenous (IV) ketamine administration. This administration route has limitations that can be overcome with oral ketamine. As such, this study undertook the first open-label low dose Oral Ketamine Trial on PTSD (OKTOP) to determine the safety and feasibility of sub-anaesthetic ketamine for PTSD symptom reduction. Participants with PTSD (n = 22 adults, aged 22-77 years, 55% female, 82% with comorbid depression) followed a weekly treatment course of low dose oral ketamine (titrated from 0.5 mg/kg to a maximum of 3.0 mg/kg) for six weeks. The primary outcome measure was the PTSD Checklist (PCL-5), with secondary measures including scales for depression, anxiety, stress, suicidality, sleep, and wellbeing. Mean PCL-5 scores were significantly reduced from a pre-ketamine baseline score of 40 to a post-ketamine score of 17 and remained at a reduced score (21) at follow-up, 1-month post-treatment. This reduction resulted in a response rate (defined as a ≥50% reduction in PCL-5 score from baseline) of 73% post-ketamine and 59% at follow-up. This response rate is comparable with IV ketamine trials for PTSD and suggests oral ketamine administration is a feasible and tolerable treatment for PTSD.

## Introduction

Up to 70% of people experience at least one traumatic event in their lifetime (Benjet et al., 2016). Whilst most people will suffer only short-term impacts from such events, ∼10% will go on to experience the prolonged symptoms characteristic of post-traumatic stress disorder (PTSD) (Australian Bureau of Statistics 2020-21). The Diagnostic and Statistical Manual of Mental Disorders (5^th^ ed) (DSM-5) categorises PTSD within the Trauma- and Stressor-Related Disorders (American Psychiatric Association, 2013). In addition to having trauma exposure, there are four symptom clusters integral and necessary to a diagnosis of PTSD: (1) re-experiencing (intrusive thoughts, flashbacks, nightmares), (2) avoidance (of contextual or geographical triggers/reminders or internal sensations (thoughts, memories, physical symptoms)), (3) increased negative affect (depression, hopelessness, difficulty concentrating), and (4) hyperarousal (heightened physiological and emotional reactivity) (American Psychiatric Association, 2013). To complete the diagnostic criteria, symptoms must be present for at least one month, not be caused by drugs or other illnesses, and cause significant functional impairment (American Psychiatric Association, 2013).

PTSD can develop from trauma exposure in various settings and have profound impacts on those affected. The highest rates of PTSD have been reported in survivors of sexual assault, military war and imprisonment, and genocidal crimes worldwide (American Psychiatric Association, 2013). Epidemiological data reveals that over 90% of individuals with PTSD experience a comorbid psychiatric disorder including suicidal ideation, anxiety, alcohol use disorders, and depression (Sareen, 2014). In fact, individuals with PTSD are 80% more likely than individuals without PTSD to meet the diagnostic criteria for at least one additional mental illness (American Psychiatric Association, 2013). Specifically, one study found 77% of women and 86% of men with PTSD also had another disorder, such as affective (51% women, 52% men), anxiety (52% men, 54% women), or substance use disorders (Mills et al., 2011). Overall, PTSD is often linked to poor social and familial relationships, job absenteeism, and lower income, educational attainment, and occupational accomplishment (American Psychiatric Association, 2013).

In contemporary clinical settings, PTSD treatment often includes a combination of psychological and pharmacological interventions (Green, 2013; Thomas and Stein, 2017). Trauma-focused psychotherapies with the largest current evidence base are exposure-based treatments and include Prolonged Exposure (PE), Cognitive Processing Therapy (CPT), trauma-focused Cognitive Behavioural Therapy (CBT), and Eye Movement Desensitization and Reprocessing (EMDR) (Norman et al., 2023; Watkins et al., 2018). Exposure therapy decreases symptoms by having patients envision themselves exposed to stimuli rather than avoiding them. By encouraging adaptive associations with painful memories, this approach improves inhibitory learning and reduces suffering, rendering the trauma tolerable (López-Ojeda and Hurley, 2022). These therapeutic interventions concentrate on the cognitions, emotions, memories, and thoughts that are linked to the traumatic events (López-Ojeda and Hurley, 2022; Watkins et al., 2018). Beyond exposure-based treatments, alternatives like Reconsolidation Therapy (RT), which focuses on how traumatic events are stored as memories (Brunet et al., 2021), are being investigated. However, further research is needed in this space to establish robust alternatives to current exposure-based psychotherapies.

Pharmacotherapy for PTSD is an option when psychotherapy has been found ineffective, individual patients have different preferences, or individuals cannot directly engage in psychotherapy (especially in the initial presentation). Standard PTSD treatment protocols suggest antidepressants, particularly selective serotonin reuptake inhibitors (SSRIs), as the first-line treatment medication for PTSD (Asnis et al., 2004). However, SSRIs are known to take up to six weeks to reduce hyperarousal, anxiety, suicidal thoughts, and depressive symptoms (Frazer and Benmansour, 2002). Two SSRIs are FDA-approved for PTSD treatment, paroxetine and sertraline, and collectively, response rates to these drugs rarely exceed 60%, with less than 30% of patients achieving full remission (Asnis et al., 2004; Berger et al., 2009). Sleep disruption, drowsiness, sexual dysfunction, weight gain, withdrawal symptoms, and decreased therapeutic response are all commonly reported with chronic SSRI use and limit their overall efficacy (Ferguson, 2001). Similar to SSRIs, selective serotonin and norepinephrine reuptake inhibitors (SNRIs) have been evaluated in PTSD, but also show limited efficacy and often leave residual symptoms (Jeffreys et al., 2012). Other drug classes, including monoamine oxidase inhibitors (MAOIs) and tricyclic antidepressants (TCAs), are sometimes considered, but have limited utility due to higher adverse effects and safety issues (Asnis et al., 2004). Finally, some small improvements have been reported with atypical antipsychotics, which can be used with antidepressants. Atypical antipsychotics can have mood stabilising properties, improve sleep patterns (decreasing the frequency of nightmares) and reduce the frequency of flashbacks and intrusive thoughts (Adetunji et al., 2005). However, they are considered primarily an adjunct to standard treatments in selectively warranted cases.

Ketamine, an N-methyl-D-aspartate-type glutamate (NMDA) receptor antagonist, has been used in medicine as an aesthetic and analgesic since the 1970s (Kurdi et al., 2014). Starting around 2000, low dose (i.e. sub-anaesthetic) ketamine began being tried off label for treatment-resistant depression (TRD), with studies in this space continuing to today (Berman et al., 2000; Ragnhildstveit et al., 2023). Although intravenous (IV) infusion is the most common route of administration, a ketamine nasal spray formulation received FDA approval in 2019 for the treatment of TRD (Kim et al., 2019). Investigations into ketamine’s effectiveness for PTSD treatment began appearing in the literature in 2005 and, to date, systematic review has identified 26 studies that retrospectively or prospectively examined ketamine use for PTSD in the form of case studies (n=7), chart reviews (n=7), open-label studies (n=5) or randomised control trials (RCTs) (n=7) (Ragnhildstveit et al., 2023). Of these PTSD ketamine studies, the vast majority have employed IV infusion as the route of ketamine administration (in four of the case studies, five of the chart reviews and all the open-label studies and RCTs) (Ragnhildstveit et al., 2023). Representing the other routes of ketamine administration, intranasal and intramuscular administration has been reported from one case study and one chart review each, while oral ketamine administration has been reported on from one case study and two chart reviews (Ragnhildstveit et al., 2023). Since that systematic review, an additional oral ketamine administration case study report has been published (Veraart et al., 2023).

Because open-label studies and RCTs have been limited to IV ketamine administration in the PTSD treatment space, there is a significant gap in our knowledge regarding the safety, tolerability, and efficacy of ketamine for PTSD delivered by non-IV routes. While IV ketamine has been found to be safe and well tolerated in open-label studies and RCTs (Ragnhildstveit et al., 2023), this method must be administered in a hospital or clinic setting under medical supervision, making it difficult to implement in conventional clinical psychiatry. This limits its use in ambulatory settings, especially for treatment programs that require daily or intermittent dosing schedules (Andrade, 2019). Alternatively, the oral administration of ketamine is more practical and the simplest route for implementation, giving it higher clinical applicability. Oral ketamine also has a much lower cost compared to other administration forms, offering a significant opportunity for financially disadvantaged cohorts (Andrade, 2019; Can et al., 2021; Muhorakeye and Biracyaza, 2021).

Collectively, the advantages of oral ketamine as a viable treatment for individuals with PTSD warrants further investigation to determine its potential efficacy. However, a substantial amount of knowledge remains deficient concerning dosage levels, treatment protocols, and the feasibility and safety of ketamine use over the short- and long-term when administered orally. To investigate this matter, the current research conducted an open-label, flexible-dose, 6-week trial of oral ketamine, followed by a 4-week follow-up phase without ketamine. The purpose of the trial was to evaluate the efficacy and safety of the medication, as well as the acute and prolonged responses, in the treatment of PTSD. The primary outcome measure was change in PTSD-related scales (PCL-5). We hypothesized that the administration of oral ketamine, once weekly for six weeks, would result in a reduction in PTSD symptomatology from a pre-ketamine baseline to a post-treatment follow-up.

## Methods

### Ethics approval and consent

Metro North Health Human Research Ethics Committee (HREC) granted approval for this study (Project ID: 42836; HREC/18/QPCH/28), and the University of the Sunshine Coast HREC subsequently confirmed that approval (A181190). The study was conducted in adherence to the ethical guidelines outlined in the Declaration of Helsinki. Every participant delivered a written consent that was well-informed. Fuel vouchers served as the form of compensation. The trial was officially documented with the Clinical Trials Registry of Australia and New Zealand (ACTRN12618001965291).

### Participants

Psychiatrists and local general practitioners (GPs) were notified of the trial through clinic visits and telephone conversations. Approved advertising materials were provided to local GP clinics and hospitals and were made available on our website. The study recruited participants who had received a diagnosis of PTSD from their general practitioners, psychiatrists, and psychologists in the area.

The study recruited individuals who were 18 years of age or older and had a confirmed diagnosis of PTSD, as determined by the Clinician Administered PTSD Scale for DSM-5 (CAPS-5) (Weathers et al., 2018). The principal investigator (a consultant psychiatrist) confirmed this through comprehensive psychiatric evaluation and mental state evaluation of the participants and the administering of a CAPS-5 assessment. Participants who were assessed for the study but were not enrolled were documented by the investigators. Eligible participants for the study were those who fulfilled all the inclusion criteria and did not satisfy any of the exclusion criteria.

Exclusion criteria were (1) Psychiatric conditions: (a) psychosis, (b) mania or hypomania, (c) acute suicidality requiring urgent psychiatric intervention and (d) a history of ketamine use disorder, (2) Physical conditions: (a) participants who have history of epilepsy or unexplained seizure history, (b) uncontrolled/severe symptomatic cardiovascular disease states including recent myocardial infarction (within prior 6 months), history of stroke and hypertension (resting blood pressure >150/100), (c) body weight of >150 kg, (d) history of intracranial mass, intracranial haemorrhage/stroke, cerebral trauma/traumatic brain injury or increased intracranial pressure (as assessed by referring general practitioner), (f) liver function test (LFT) results out of normal range, (g) previous reaction to ketamine (as reported by referring general practitioner and participant), (h) participants who are pregnant, currently breastfeeding, or who are planning a pregnancy during the trial, (i) participants who are simultaneously engaging in another clinical intervention, (j) participants with a history of substance use disorder (excluding ketamine use disorder) may be eligible to participate in the study if they abstain from alcohol or illicit substance use two weeks prior to participation in the trial and for the remainder of the trial.

The physical health of all participants was evaluated through a comprehensive assessment that included physical examination, laboratory blood testing, and a review of their medical history conducted by the primary investigator/psychiatrist. Throughout the treatment and follow-up phase, participants continued their regular prescribed medication as directed by their respective healthcare providers. Most individuals in the study were concurrently taking psychopharmacological drugs, such as SSRIs, SNRIs, antipsychotics and mood stabilisers (**Table 1**).

**Table 1.**
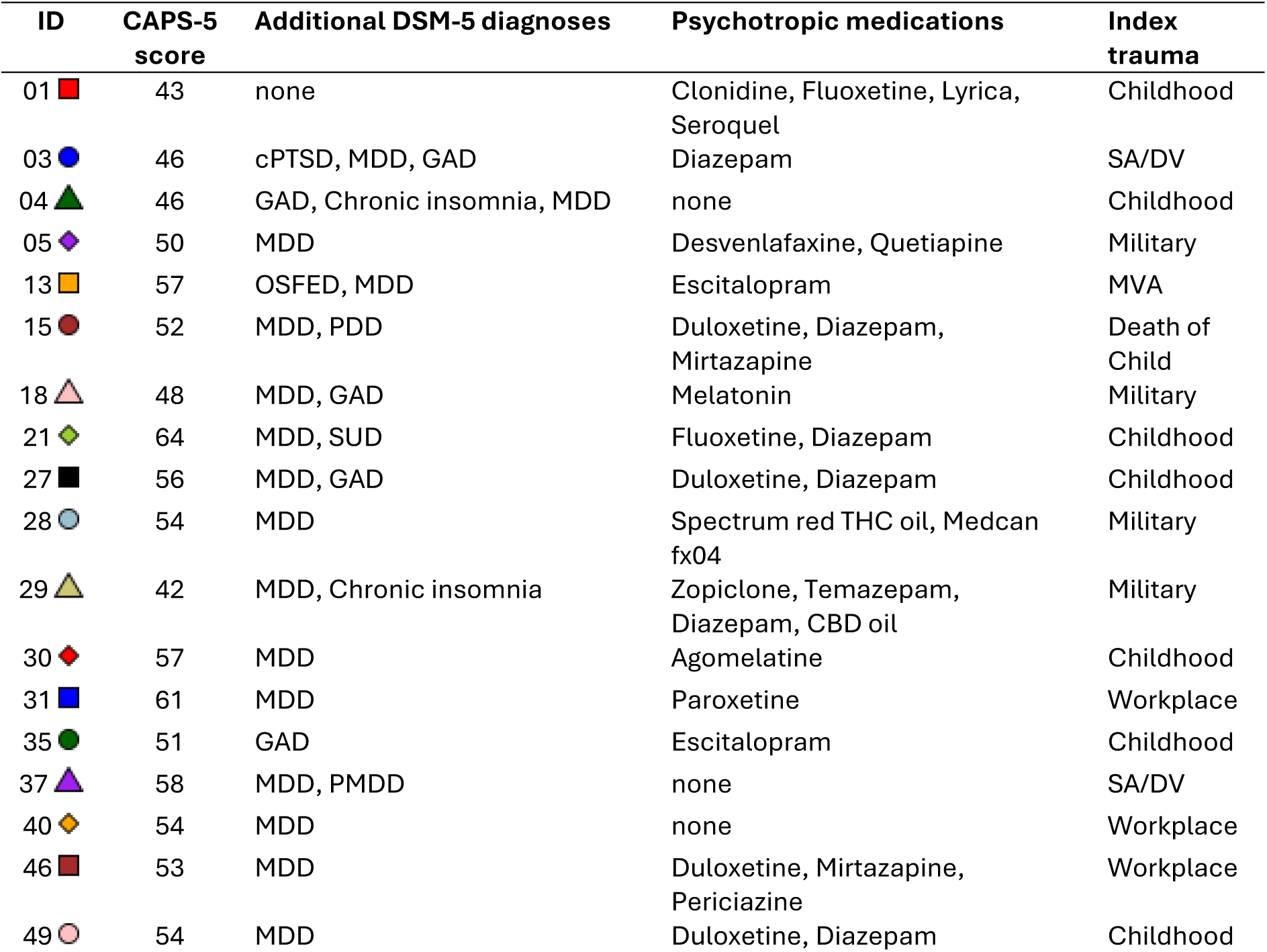
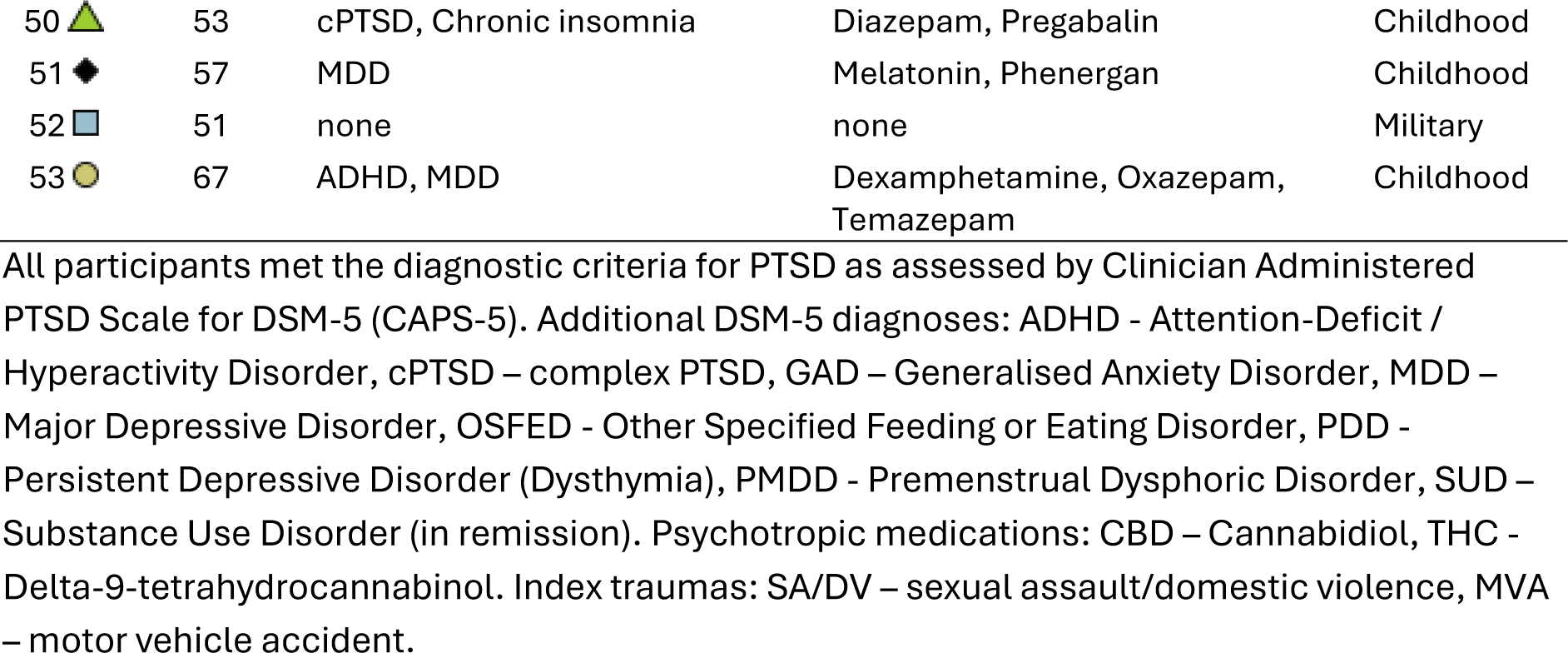
CAPS-5 scores, co-morbidities, concomitant psychotropic medications, and index trauma of study participants at the baseline assessment.

| ID | CAPS-5 score | Additional DSM-5 diagnoses | Psychotropic medications | Index trauma |
| --- | --- | --- | --- | --- |
| 01 | 43 | none | Clonidine, Fluoxetine, Lyrica, Seroquel | Childhood |
| 03 | 46 | cPTSD, MDD, GAD | Diazepam | SA/DV |
| 04 | 46 | GAD, Chronic insomnia, MDD | none | Childhood |
| 05 | 50 | MDD | Desvenlafaxine, Quetiapine | Military |
| 13 | 57 | OSFED, MDD | Escitalopram | MVA |
| 15 | 52 | MDD, PDD | Duloxetine, Diazepam, Mirtazapine | Death of Child |
| 18 | 48 | MDD, GAD | Melatonin | Military |
| 21 | 64 | MDD, SUD | Fluoxetine, Diazepam | Childhood |
| 27 | 56 | MDD, GAD | Duloxetine, Diazepam | Childhood |
| 28 | 54 | MDD | Spectrum red THC oil, Medcan fx04 | Military |
| 29 | 42 | MDD, Chronic insomnia | Zopiclone, Temazepam, Diazepam, CBD oil | Military |
| 30 | 57 | MDD | Agomelatine | Childhood |
| 31 | 61 | MDD | Paroxetine | Workplace |
| 35 | 51 | GAD | Escitalopram | Childhood |
| 37 | 58 | MDD, PMDD | none | SA/DV |
| 40 | 54 | MDD | none | Workplace |
| 46 | 53 | MDD | Duloxetine, Mirtazapine, Periciazine | Workplace |
| 49 | 54 | MDD | Duloxetine, Diazepam | Childhood |
| 50 ▲ | 53 | cPTSD, Chronic insomnia | Diazepam, Pregabalin | Childhood |
| 51 ◆ | 57 | MDD | Melatonin, Phenergan | Childhood |
| 52 ■ | 51 | none | none | Military |
| 53 ● | 67 | ADHD, MDD | Dexamphetamine, Oxazepam, Temazepam | Childhood |

All participants met the diagnostic criteria for PTSD as assessed by Clinician Administered PTSD Scale for DSM-5 (CAPS-5). Additional DSM-5 diagnoses: ADHD - Attention-Deficit / Hyperactivity Disorder, cPTSD – complex PTSD, GAD – Generalised Anxiety Disorder, MDD – Major Depressive Disorder, OSFED - Other Specified Feeding or Eating Disorder, PDD - Persistent Depressive Disorder (Dysthymia), PMDD - Premenstrual Dysphoric Disorder, SUD – Substance Use Disorder (in remission). Psychotropic medications: CBD – Cannabidiol, THC - Delta-9-tetrahydrocannabinol. Index traumas: SA/DV – sexual assault/domestic violence, MVA – motor vehicle accident.

Following the provision of informed consent, a psychiatric review was conducted by the psychiatrist and study nurses in the form of a semi-structured interviews to elicit a diagnosis of PTSD and any comorbidities (**Table 1**), and subsequent eligibility for the study, including all wellbeing and safety scales. Data pertaining to demographic details, concomitant pharmaceutical treatment and medical conditions were also collected. Vital signs, blood tests and urinalysis were taken to establish baselines and confirm the absence of contraindications (e.g., uncontrolled hypertension and pregnancy). A registered nurse undertook vital signs and urinalysis.

### Study design and treatment

From August 2021 to January 2024, this open-label trial of oral ketamine was conducted at the Thompson Institute, University of the Sunshine Coast. A 6-week flexible-dose course of oral (racemic) ketamine was administered for the treatment for PTSD, followed by a 4-week follow-up phase (i.e., without ketamine) to evaluate the dosing regimen, safety/tolerance, and acute and prolonged response rates to once-weekly oral ketamine administration.

In the fourteen days prior to the commencement of ketamine treatment, all participants underwent a baseline assessment that included a physical examination, urinalysis, laboratory blood testing, and review of their medical history. Over a 6-week treatment period, each participant was administered one dose of ketamine per week as an oral subanesthetic. Ketamine was prescribed by the consultant psychiatrist and administered as a liquid in fruit juice at an initial dosage of 0.5 mg/kg by the registered nurse or nurse practitioner. Depending on patient tolerance, dose amounts were increased by 0.1–0.5 mg/kg at each treatment, to a maximal dose of 3.0 mg/kg at the sixth treatment. The assessment of urinalysis and the presence or absence of adverse effects, as determined by tolerability and safety rating instruments, was employed to ascertain ketamine intolerance. Participants were under immediate post-treatment supervision for a minimum of sixty minutes. For twelve hours post-treatment, participants were instructed to refrain from engaging in activities that demanded alertness (such as operating a motor vehicle), adhering to work, or entering legal contracts. On the day of the treatment, they were also required to arrange for appropriate transportation to and from the location (not allowed to drive themselves). The overall trial design is illustrated in **Figure 1**, which highlights three significant time points: (i) “pre-ketamine” (up to −2 weeks prior to first treatment), which comprised the “baseline” timepoint; (ii) “post-ketamine”, which occurred 1–7 days after the final treatment, and (iii) “follow-up,” which occurred 28–32 days after the final treatment, respectively. Phone evaluations were conducted 24-hours post-treatment to monitor for adverse effects.

**Figure 1.**
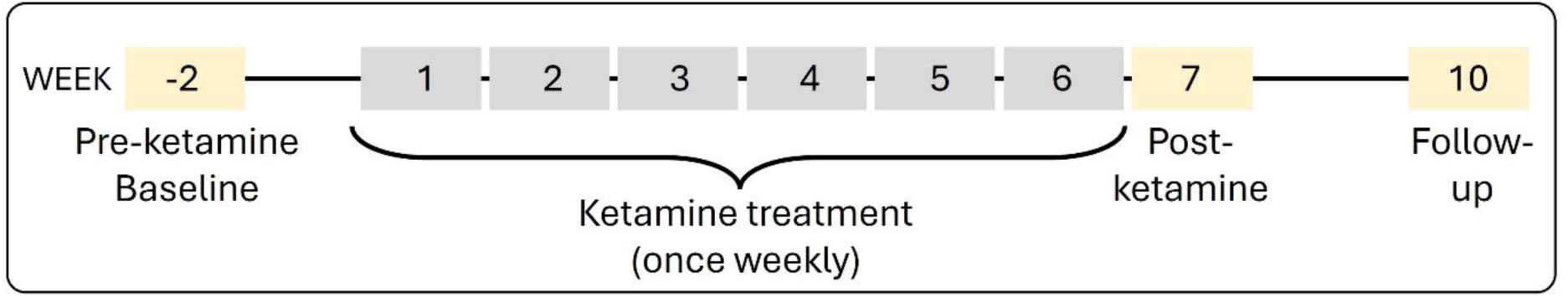
Oral Ketamine Trial on PTSD (OKTOP) study design. Following written consent and eligibility confirmation, OKTOP participants underwent a pre-ketamine baseline assessment which included physical examination, urinalysis, laboratory blood testing, review of medical history, full clinical scales assessment (including CAPS-5 for baseline only), research blood collection, magnetic resonance imaging (MRI) and electroencephalogram (EEG) imaging, and cognitive functioning assessment. During ketamine treatment, weekly assessments included physical examination, urinalysis, and full clinical scales assessment, with treatment weeks 3 and 6 also collecting research bloods and MRI & EEG imaging. Post-ketamine and Follow-up assessments included full clinical scales assessment, laboratory blood testing, research blood collection, MRI & EEG imaging, and cognitive functioning assessment. This report focuses on the clinical scale assessment outcomes only.

### Assessments

Physical assessments were conducted at each study visit to monitor participant health and determine adverse events. At pre-ketamine baseline and prior to all treatments, urinalysis, using a bedside test, was performed by clinical staff to screen for cystitis. Urine pregnancy screening was undertaken for females of childbearing age at the pre- and post-ketamine time points, as well as pregnancy tests as required. At each treatment visit, vital signs (heart rate, blood pressure, oxygen saturation level, and temperature) were recorded three times: before ketamine delivery and at 30- and 60-minutes post-treatment. Standard clinical pathology laboratory blood testing was performed at pre-ketamine baseline, post-ketamine and follow-up and included Full Blood Count (FBC), Urea and Electrolytes (UE), LFT, and Thyroid Function Test (TFT).

In addition to the CAPS-5 assessment (Weathers et al., 2018) conducted at pre-ketamine baseline, the following clinical rating scales were completed by participants at baseline, post-ketamine and follow-up timepoints to evaluate the primary and secondary trial outcomes: (a) PTSD Checklist for DSM-5 (PCL-5), a 20-item self-report measure often used in conjunction with the CAPS-5 to monitor PTSD symptom change across patient treatment (Blevins et al., 2015; Weathers et al., 2018); (b) Life Events Checklist for DSM-5 (LEC-5), a 17-item self-report measure designed to evaluate participant’s exposure to potentially traumatic events across the lifetime, often used in conjunction with CAPS-5 (baseline only) (Elhai et al., 2005); (c) Beck Scale for Suicide Ideation (BSS), a 21-item clinician-rated scale which examines suicidal intent in patients (Beck et al., 1979); (d) Montgomery-Asberg Depression Rating Scale (MADRS), a 10-item clinician-rated scale used to measure the severity of depressive symptoms (Montgomery and Asberg, 1979); (e) Depression, Anxiety, and Stress Scale (DASS-21), 21-item self-report measure assessing perceived depressive, anxiety, and stress symptoms (Lovibond and Lovibond, 1995); (f) Social and Occupational Functioning Assessment Scale (SOFAS), a clinician-rated single-item scale used to indicate an individual’s level of social and occupational functioning (Goldman et al., 1992); (g) World Health Organization Well-Being Index (WHO-5), a 5-item self-report global rating scale measuring subjective well-being (Bech et al., 2003); (h) Pittsburgh Sleep Quality Index –Addendum for PTSD (PSQI-A), a 7-item self-report measure assessing frequency of disruptive nocturnal behaviours specific to PTSD (Germain et al., 2005); (i) Perceived Stress Scale (PSS), 10-item self-report measure assessing a patient’s perception of stress (Cohen et al., 1983); (j) Snaith-Hamilton Pleasure Scale (SHAPS), 14-item clinician-rated scale evaluating anhedonia for neuropsychiatric disorders (Franken et al., 2007).

The following safety and tolerability measures were also collected 60 min after each ketamine dose in-person and 24-hours post-treatment via a phone call: (a) Patient Rated Inventory of Side Effects (PRISE), 33-item self-report measure assessing side-effects within specific symptom domains across the past 7 days (Rush et al., 2004); (b) Clinician Administered Dissociative States Scale (CADSS), a structured clinical interview used to check for present-state dissociative symptoms (Bremner et al., 1998); (c) Brief Psychiatric Rating Scale (BPRS), an 18-item clinician-rated scale used as a general measure of psychiatric symptoms such as depression, anxiety, hallucinations and unusual behaviour (Overall and Gorham, 1988); (d) Young Mania Rating Scale (YMRS), a 11-item clinician-rated scale used to check for elevated mood (i.e. mania) (Young et al., 1978). CADSS, BPRS and YMRS items relating to positive symptoms were closely monitored for potential side effects.

### Outcome measures

The primary outcome measure for the study was a reduction in PTSD symptomology with ketamine treatment, as determined by the PCL-5 scale. A ‘response’ was defined as a ⩾50% improvement in PCL-5 score from the pre-ketamine to the post-ketamine visit. A ‘prolonged response’ was defined as a ⩾50% improvement in PCL-5 score from pre-ketamine to the follow-up visit. Secondary outcome measures included pre-ketamine to follow-up changes in BSS, DASS-21 (as the three individual subscales of anxiety, depression, and stress), MADRS, SOFAS, and WHO-5 scores. Additionally, PSQI-A, PSS and SHAPS-C scores were also evaluated pre-ketamine to follow-up.

### Analysis

Statistical analyses were carried out using SPSS Statistics for Windows, version 27 (IBM Corp, Armonk, NY). Assumptions were checked for all analyses and reported in results. To test the primary outcome, a repeated measures ANOVA was conducted with time as the within subjects’ factor (three levels: pre-ketamine, post-ketamine and follow-up timepoints) and PCL-5 as the dependent variable. Pairwise comparisons of the timepoints were performed with Bonferroni corrections. Cohen’s d effect sizes were also calculated to further describe differences between timepoints. Repeat measure ANOVAs were additionally conducted for secondary outcomes, BSS, MADRS, DASS, SOFAS and WHO-5, as well as PSQI-A, PSS and SHAPS-C, as described above, with three levels of time. For all tests, significance thresholds were set at p < 0.05.

## Results

Of the 131 participants pre-screened for this study, n=59 met initial eligibility and n=38 were enrolled in the study. Of these, six participants failed to attend the baseline assessment and subsequently withdrew, and five participants failed to meet final eligibility (did not meet CAPS-5 requirement or medical criteria during the baseline assessment). Of the remaining 27 participants, five participants missed more than one treatment week and/or the post-treatment or follow-up assessment and were removed from analysis. This resulted in a final study cohort of n=22 participants that met all eligibility criteria and completed all required treatments and assessments.

The study cohort (12 female, 10 male) had a mean age of 47.07 ± 13.76 years (range 22.39-77.93 years) and a mean weight at first treatment of 90.85 ± 23.52 kg. In addition to PTSD, n=18 (82%) met DSM-5 criteria for comorbid major depressive disorder (MDD) and n=5 (26%) met criteria for comorbid generalised anxiety disorder (GAD). Additionally, n=5 (23%) were medication-free at trial commencement. The Index Event trauma reported varied for participants and included childhood trauma (n=10, 45%), military trauma (n=5, 23%), workplace trauma (n=3, 14%), sexual assault/domestic violence (n=2, 9%), motor vehicle accident (n=1, 5%) and death of a child (n=1, 5%) (**Table 1**).

### Primary outcome measure

The mean PCL-5 score was significantly reduced at both post-ketamine (16.96 ± 16.18, n=22; range=0–57) and follow-up (21.05 ± 16.62, n=22; range=0–63) timepoints (**Figure 2**) when compared to the pre-ketamine timepoint (40.32 ± 11.61, n=22; range=12–61). This was confirmed by omnibus ANOVA (main effect of time; p <0.001) and significant pairwise comparisons between pre-ketamine and post-ketamine (p <0.001; d=1.30) and follow-up (p <0.001; d=1.05) timepoints, respectively. There was a small increase in the PCL-5 score from post-ketamine to follow-up, however this was not significant (pairwise comparison, p=0.110; d=−0.47). Thus, at the group level, the PCL-5 score reduced to below the clinical threshold score of 31 at the post-ketamine timepoint. Furthermore, the mean follow-up PCL-5 score remained at almost half of the mean pre-ketamine PCL-5 score.

**Figure 2.**
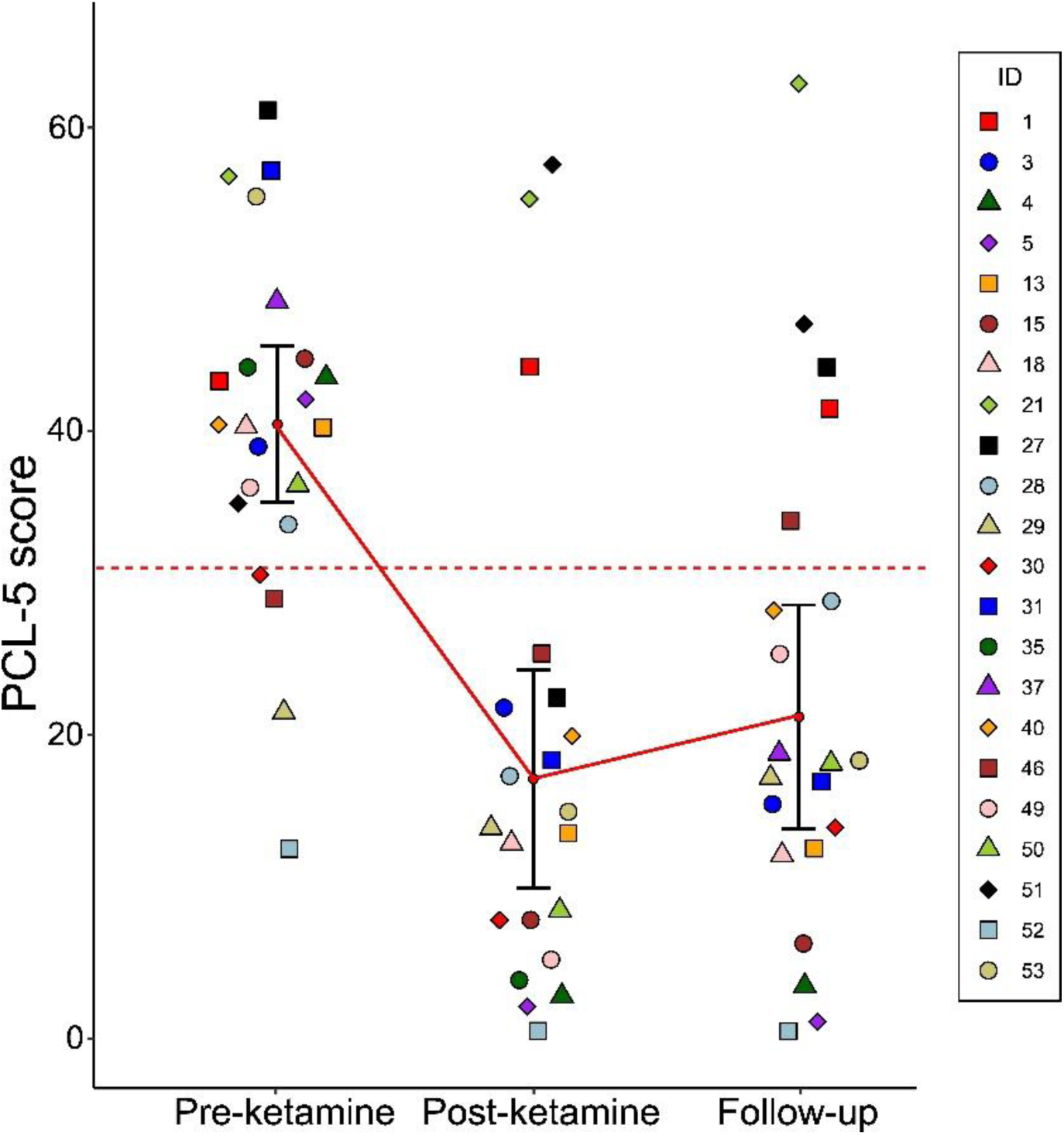
PTSD Checklist for DSM-5 (PCL-5) scores at major study timepoints. Points represent individual participant PCL-5 scores at Pre-ketamine (baseline), Post-ketamine (1-week post-treatment), and Follow-up (1-month post-treatment). Dashed red line signifies threshold (>31) indicative of a PTSD diagnosis by PCL-5. Mean and 95% confident interval (CI) are indicated by the red circle and error bars, respectively. (Note – all participants met criteria for a clinical diagnosis of PTSD by CAPS-5 at Pre-ketamine.)

The proportion of PCL-5 responders (participants with ≥50% reduction in PCL-5 score) was 73% at post-treatment and remained at 59% at follow-up (prolonged responders). Notably, n=12 participants were responders at both post-ketamine and follow-up timepoints.

### Secondary outcome measures

The secondary outcome measures, also collected at pre-ketamine, post-ketamine, and follow-up timepoints, all showed significant reductions (**Table 2**). Specifically, the BSS scores reduced from 5.68 pre-ketamine to 1.73 post-ketamine and 2.36 at follow-up (p<0.001) and, while the mean score for this scale was below the clinical threshold at baseline, this reduction represented a significant lowering of suicidality risk in this cohort. Additionally, the MADRS score decreased from 32.77 at pre-ketamine (severe depression range) to 8.55 post-ketamine and 9.91 at follow-up (mild depression range). The self-rated DASS scale showed significant reductions from pre-ketamine to post-ketamine and follow-up across all 3 subscales (p<0.001). Notably, DASS scores at the post-ketamine timepoint were all in the ‘Normal’ range, with anxiety and stress subscales remaining ‘Normal’ at follow-up, and the depression subscale elevating to only the ‘Mild’ range. SOFAS scores, indicative of social and vocational functioning, significantly improved from 59.55 pre-ketamine to 76.82 post-ketamine. The self-rated WHO-5 showed a highly significant improvement in quality of life, from 28.55% at pre-ketamine to 54.55% post-ketamine (above 50% threshold for depression) and dipping slightly to 49.64% at follow-up. Finally, additional measures for sleep (PSQI-A), perceived stress (PSS) and pleasure (SHAPS) all showed significant improvements at post-ketamine and follow-up (**Table 2**).

**Table 2.**
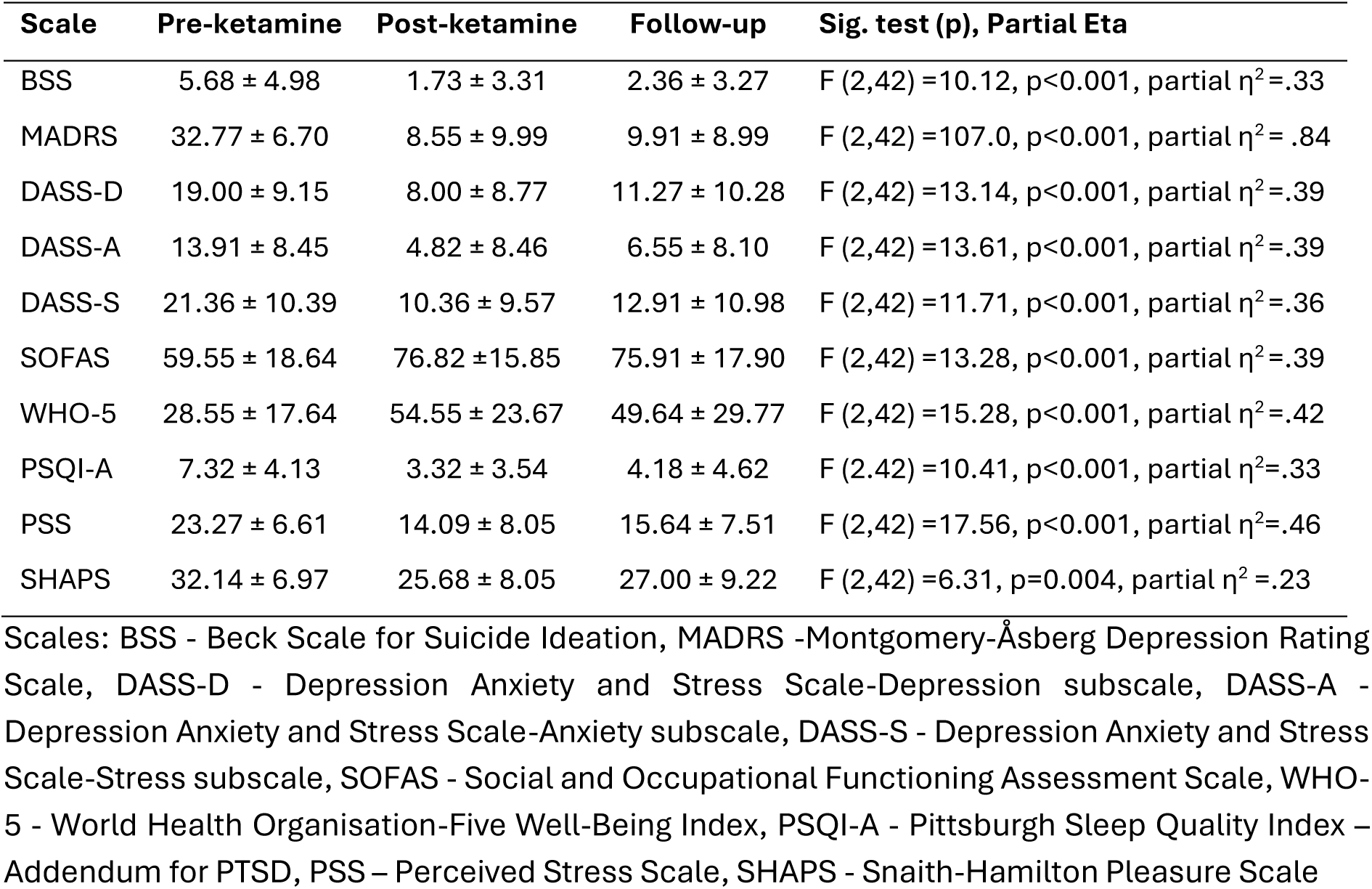
Mean scores (± standard deviation) for secondary outcomes at pre-ketamine, post-ketamine and follow up time-points for N=22.

### Adverse events and side effects

No serious adverse events were recorded during the study. Transient changes in vital signs were observed throughout treatment but no significant variations in heart rate, respiratory rate or oxygen saturation were observed. The treatment was generally well tolerated, with no participants withdrawing from the trial due to adverse side effects due to the ketamine.

The most frequent side effects reported throughout the six weeks of ketamine treatment were difficulty sleeping, headache, anxiety, fatigue, poor concentration, decreased energy, followed by restlessness, general malaise, dry mouth, dizziness, and dry skin (**Table 3**). These side effects were consistent with those observed in a similar low dose oral ketamine trial for suicidality (Can et al., 2021). Reports of distressing side effects decreased from 4% of total symptoms monitored at treatment 1 to between 1-2% for treatments 2-6 (**Table 3**).

**Table 3.** Adverse symptoms from Patient Rated Inventory of Side Effects (PRISE).

| Symptoms | Tx1 | Tx2 | Tx3 | Tx4 | Tx5 | Tx6 |
| --- | --- | --- | --- | --- | --- | --- |
| **Gastrointestinal symptoms** |  |  |  |  |  |  |
| Diarrhea | 3/1 | 1/0 | 4/0 | 4/0 | 1/0 | 4/0 |
| Constipation | 1/0 | 1/1 | 1/1 | 0/0 | 1/0 | 3/0 |
| Dry mouth | 7/1 | 9/0 | 5/0 | 7/1 | 8/0 | 8/0 |
| Nausea/vomiting | 2/0 | 4/0 | 0/0 | 4/0 | 5/0 | 3/0 |
| **Heart symptoms** |  |  |  |  |  |  |
| Palpitation (skipping a beat) | 0/0 | 0/0 | 0/0 | 2/0 | 1/0 | 3/0 |
| Dizziness on standing | 3/0 | 2/0 | 2/0 | 3/0 | 3/0 | 3/0 |
| Chest pain | 0/0 | 0/0 | 0/0 | 1/0 | 0/0 | 2/0 |
| **Skin symptoms** |  |  |  |  |  |  |
| Rash | 0/0 | 0/0 | 1/0 | 0/0 | 0/0 | 0/0 |
| Increased perspiration | 3/1 | 1/1 | 1/0 | 2/0 | 3/0 | 0/1 |
| Itching | 1/0 | 2/0 | 1/0 | 3/0 | 2/0 | 4/0 |
| Dry skin | 5/1 | 3/1 | 2/1 | 2/1 | 3/1 | 5/1 |
| **Nervous system symptoms** |  |  |  |  |  |  |
| Headache | 11/1 | 4/0 | 7/1 | 7/2 | 6/1 | 8/0 |
| Tremors | 0/0 | 2/0 | 0/0 | 2/0 | 2/0 | 2/0 |
| Poor coordination | 2/0 | 1/0 | 1/0 | 0/0 | 2/0 | 1/0 |
| Dizziness | 4/0 | 4/0 | 2/0 | 5/0 | 4/0 | 3/0 |
| **Eyes / ears symptoms** |  |  |  |  |  |  |
| Blurred vision | 1/1 | 1/1 | 1/0 | 1/0 | 1/1 | 2/1 |
| Ringing in ears | 3/0 | 5/0 | 4/0 | 3/1 | 3/1 | 5/0 |
| **Urinary / menstrual symptoms** |  |  |  |  |  |  |
| Difficulty urinating | 0/0 | 0/0 | 0/0 | 0/0 | 0/0 | 0/0 |
| Painful urination | 0/0 | 0/0 | 0/0 | 0/0 | 0/0 | 0/0 |
| Frequent urination | 2/0 | 1/0 | 3/0 | 3/0 | 3/0 | 3/0 |
| Menstrual irregularity | 1/1 | 1/0 | 2/0 | 2/0 | 2/0 | 0/0 |
| **Sleep symptoms** |  |  |  |  |  |  |
| Difficulty sleeping | 5/5 | 7/2 | 7/2 | 9/2 | 5/2 | 10/1 |
| Sleeping too much | 0/1 | 0/1 | 1/1 | 2/0 | 0/1 | 1/1 |
| **Sexual functioning symptoms** |  |  |  |  |  |  |
| Loss of sexual desire | 2/0 | 2/0 | 2/0 | 1/0 | 1/0 | 0/0 |
| Trouble achieving orgasm | 1/0 | 2/0 | 1/0 | 1/0 | 1/0 | 0/0 |
| Trouble with erections | 0/0 | 0/0 | 0/0 | 0/0 | 0/0 | 0/0 |
| **Other symptoms** |  |  |  |  |  |  |
| Anxiety | 8/4 | 9/0 | 12/0 | 11/0 | 8/0 | 9/0 |
| Fatigue | 6/4 | 8/2 | 8/2 | 7/2 | 5/3 | 7/1 |
| Poor concentration | 7/3 | 7/1 | 8/1 | 8/0 | 8/2 | 7/0 |
| Decreased energy | 6/2 | 9/1 | 8/1 | 8/0 | 5/1 | 5/1 |
| General malaise (discomfort) | 1/0 | 3/1 | 4/0 | 6/0 | 3/0 | 2/0 |
| Restlessness | 6/1 | 7/0 | 7/0 | 9/0 | 5/0 | 6/0 |

| Other | 4/2 | 1/0 | 1/0 | 0/1 | 1/1 | 1/0 |
| --- | --- | --- | --- | --- | --- | --- |
| Reported as N(tolerable)/N(distressing) from n = 22 participants at 24 hr post-ketamine treatment weekly. |  |  |  |  |  |  |

Dissociation symptoms were monitored via the CADSS clinical interview 60 min post-treatment throughout the treatment schedule, as ketamine dosage increased each week based on patient tolerance. At treatment 1 (with a dosage of 0.5 mg/kg), six participants (27%) reported mild effects (“I feel slightly detached from what is going on, but I am basically here”), while only one participant reported moderate effects (“Things seem dreamlike, although I know I am awake”). As treatments continued, dissociation effects remained limited, with most effects reported as mild or moderate. Severe dissociation was only reported from one participant at treatment 4 and two participants at treatment 5 and 6, while extreme dissociation was only reported from one participant at treatment 5. These effects dissipated quickly and did not impact the treatment schedule.

BPRS positive symptoms (delusion, conceptual disorganisation, hallucination, excitement, grandiosity, suspiciousness, hostility) were monitored 60 min post-treatment. Only two participants returned a rating above mild at any point in the study (one at treatment 1 (elevated suspiciousness and hostility) and one at treatment 3 (elevated hostility), however, re-evaluation at 24-hour post-treatment determined that these symptoms had subsided.

Finally, YMRS scores were all below the threshold for mania, with only one participant having a score of 5 the day after treatment 5 (due to less sleep and irritability) and the remainder of participants scoring 4 or below throughout the treatment period.

## Discussion

To the best of our knowledge, this study is the first to explore the feasibility, safety, and tolerability of low dose oral ketamine in an open-label clinical trial for PTSD treatment. In this pilot study, six weeks of oral ketamine (administered once a week in liquid form as a drink), led to significant decreases in PCL-5 scores (the primary outcome measure) from pre-ketamine baseline to post-ketamine and follow-up assessments. This represented 73% of participants being classified as responders to the oral ketamine treatment (≥50% reduction in PCL-5 score at the 1-week post-treatment assessment) and 59% of participants going on to be prolonged responders (maintained that ≥50% reduction in PCL-5 score at the 1-month follow-up assessment). These results are comparable to current IV ketamine trial outcomes for PTSD treatment (Ragnhildstveit et al., 2023).

In addition to significant improvements in PTSD symptomology (captured by the PCL-5), several other self-reported and clinician-rated scales also showed significant improvements with low dose oral ketamine treatment. This is an important finding, as over three-quarters of individuals diagnosed with PTSD are found to meet the criteria for at least one additional mental illness (American Psychiatric Association, 2013). In this study, low dose oral ketamine was seen to significantly improve co-morbid symptoms related to depression (via MADRS and DASS-D), suicidal ideation (via BSS), anxiety (via DASS-A), and stress (via DASS-S and PSS), as well as improve social and occupational functioning (via SOFAS), sleep (via PSQI-A) and overall pleasure and wellbeing (via SHAPS and WHO-5). These co-morbidities or functional impairments add a substantial burden to individuals living with PTSD. The finding that ketamine can improve this spectrum of symptoms in a short period of time is promising for future trials to determine efficacy and treatment strategies.

A deliberate and important aspect to this open-label pilot trial was the conservative and stepped protocol designed to gradually increase the oral ketamine dosages administrated over the treatment period. Determining the minimum dose of a medication that is both effective and tolerable involves striking a balance between minimising side effects and maximising the achievable efficacy. Despite the extensive body of research examining the impact of ketamine on diverse psychiatric disorders (Sanacora and Katz, 2018), the dose–response relationship and optimal drug administration route for ketamine remains inadequately understood. The approach employed in this study was conservative in both the maximum dosage administered and the frequency of dosage. The upper limit for oral ketamine dosage was set at 3 mg/kg, which differs from earlier research findings that have reported dosages as high as 7 mg/kg (Hartberg et al., 2018). Moreover, previous research has described more frequent dosing schedules with oral ketamine (from once every 3 days to multiple doses per day) (Al Shirawi et al., 2017; Iglewicz et al., 2015), which is in contrast to the frequency of administration in this study (once per week). By adopting an initial lower dose, this protocol was designed to assess each patient’s tolerance to oral ketamine before gradually increasing it. This strategy has been proven successful in this type of study in adults with TRD and chronic suicidality previously (Can et al., 2021). As a result, the administration of low dose oral ketamine was well received and tolerated by study participants, with the majority of reported side effects being mild and transient. Importantly, all reported side effects from oral ketamine were resolved prior to a participant’s discharge from the study. Comparatively, IV administration of ketamine has been correlated with a higher incidence of adverse effects compared to oral administration, and have encompassed physical manifestations such as headaches, dizziness, alterations in vital signs, as well as acute mental manifestations such as dissociation, anxiety, and agitation (Short et al., 2018). Additional study in the oral ketamine space has indicated that the extended-release formulation of oral ketamine also offers enhanced tolerability in relation to physical symptoms and dissociations (Glue et al., 2020). These results offer a solid foundation to continue exploring oral ketamine as a safe, well-tolerated form of ketamine treatment.

There were several notable limitations associated with this study that need to be acknowledged. Firstly, the study sample size (n=22) was relatively small and an assessment of the findings must be conducted with this in mind. Secondly, the study was open-label, with neither a placebo/active control nor a comparator study group for the oral ketamine treatment group, which limits the generalisability of the findings to the population level. Thirdly, the participants were permitted to continue with their existing treatment programs (both psychological interventions and psychopharmacological medications), and oral ketamine was administrated to participants as an augmentation. Given this, it is difficult to disentangle the contribution of the ongoing, concurrent medications/treatments. When viewed from this angle, one cannot rule out the possibility of combined or synergic effects brought about by the interaction of ketamine with other treatments. However, despite these limitations, this study represents a “real-world” trial of how oral ketamine could be integrated into the ongoing treatment programs of patients with PTSD and demonstrates that this treatment approach is feasible and well-tolerated in this population.

In contrast to conventional antidepressants (which focus primarily on monoaminergic neurotransmitters and have slow-acting effects), ketamine (via its rapid-acting glutamatergic effects) offers an important and viable treatment option for symptoms of PTSD. In this respect, this open-label clinical trial has demonstrated that low dose oral ketamine is feasible, safe, and tolerable in adults with PTSD. More research is now required in the form of larger, randomised and blinded trials to elucidate oral ketamine’s optimal dosage levels, treatment timing/protocol, and further characterise short- and long-term safety in PTSD treatment. With clinical outcomes comparable to IV infusion studies, this present study has demonstrated that low dose oral ketamine is a safe option for the treatment of PTSD and has contributed important additional information to the growing research evidence for ketamine in mental health treatment.

## Data Availability

All data produced in the present study are available upon reasonable request to the authors

## Acknowledgement

The authors express their sincere gratitude to the participants of the OKTOP study. The funding for this project was provided by the internal budget of the Thompson Institute. The authors declare that they have no conflict of interest.

